# heaterBack to work MD - Experiences of internal medicine physicians returning to work after parental leave

**DOI:** 10.64898/2026.10.06.26364854

**Authors:** Salwa Hasan, Grace H. Tang, Heather VanderMeulen, Tamara Van Bakel, Michael Fralick, Michelle Sholzberg

## Abstract

**Background:** Parenthood in medicine is an experience fraught with challenges that begin in the pre-conception phase and persist as physician-parents navigate their dual role.

**Methods:** This study aims to understand the experiences of physician-parents (both childbearing and non-childbearing) returning to work after parental leave. Physicians, including residents, affiliated with the University of Toronto’s Department of Medicine internal medicine division who had a child within the last ten years were invited to participate in a self-administered electronic survey from June2022 to 30June 2023. Data were summarized using descriptive statistics.

**Results:** Twenty-nine respondents took part in the survey; 79% were staff physicians while the remaining were trainees at the time of the survey. At the time their first child was born, 48% were staff members and 52% were trainees. The median duration of parental leave was 5 months (range 1-12 months). Among those for whom lactation was applicable, 76% reported that they continued their lactation efforts on returning to work, while 24% chose not to. Thirty five percent (35%, *n*=6/17) of respondents cited that they were not provided with duty hour modifications when required. Many respondents (46%) felt guilty about burdening their co-workers for requesting accommodations for childcare. When requested to rate their experiences of balancing their personal and professional lives on a scale of 1-10 (1 being “impossible to maintain” and 10 being “extremely easy to maintain”), most respondents (66%) chose a rating of 5 or less.

**Conclusion:** This study supports previous findings that physician-parents face a myriad of challenges integrating their roles in medicine and parenthood. A change in culture is required to support and normalize parenthood for physicians and foster sustainability and productivity in academic medicine.

## INTRODUCTION

Inclusivity statement (*adapted*^*1*^): Throughout this article, we often use the term “women” to highlight the experiences of female physicians who are mothers. We acknowledge that these terms are exclusive, and we ask the reader to please recognize that these experiences may also apply to all people with the anatomy that allows for menstruation, pregnancy, and childbirth, including girls, transgender men, intersex people, and gender nonbinary individuals.

Parenthood in medicine is an experience fraught with challenges that begin in the pre-conception phase and persist as physician-parents navigate their dual role. For example, female physicians are known to delay childbearing compared with the general population, and thus are at an increased risk of adverse reproductive outcomes including infertility and obstetrical complications^2,3^. Expectant physicians, especially those in training, must cope with physical and psychological effects of pregnancy in addition to rigorous workloads including overnight call shifts^4^. The literature describes a variety of stressors faced by physicians once their child is born, such as maintaining lactation^5,6^ and arranging reliable childcare^7,8^. Overall, motherhood in medicine has been linked to hindered career advancement^9,10^, financial inequity^11,12^ and a challenging work-life balance^9,13,14^.

This study aimed to explore the experiences of physician-parents on their return to work after parental leave among staff and trainees of the Department of Medicine (DoM) at the University of Toronto (UofT). We specifically aimed to evaluate experiences within the Canadian context, of non-surgeon physicians and of both childbearing and non-childbearing parents, as these represent gaps in the existing literature. Here, we present findings from a self-administered survey to identify barriers to successful transition back to the medical workplace along with proposed solutions to facilitate policy and programmatic change.

## MATERIALS AND METHODS

Trainee and staff physicians affiliated with Department of Medicine (DoM) at the University of Toronto (UofT) were invited to participate in an electronic survey, if they had taken at least one parental leave prior to returning to work in the 10 years preceding recruitment. Study recruitment was carried out through three separate advertisements on the DoM newsletter and approximately twenty (20) emails to personal contacts known to the study team. Recruitment occurred on a rolling basis from 10/06/2022 to 30/06/2023. Participants completed a form to indicate their consent to participate. Data was de-identified prior to the principal investigator’s review, given the potential of a reporting relationship between them and the participants. While participation was voluntary, respondents could opt in (through a separate unlinked survey) to collect a gift card as a token of appreciation. The study was approved by the Unity Health Toronto Research Ethics Board in accordance with the Tri-Council Policy Statement Ethical Conduct for Research Involving Humans and ICH Good Clinical Practice Guidelines.

The survey was composed of multiple-choice questions, with the option to choose more than one answer where applicable. Survey questions related to demographics, length of parental leave, lactation, childcare and other supports in the workplace, co-worker perceptions, and work-life integration. Open-ended questions were also included, to complement the quantitative survey data. Questions were developed following a literature review based on MEDLINE searches performed in March 2022 combining the keywords parenting, pregnancy, maternity leave, physician, and resident. Lived experiences of the researchers (S.H./M.S.) also guided survey development. The survey data were analyzed using descriptive statistics (i.e., proportions, medians, and ranges), while a selection of open-ended responses were reported verbatim.

## RESULTS

A total of 29 respondents completed the survey. Table 1 provides an overview of the demographic characteristics of the participants.

**Table 1.** Baseline characteristics of study participants.

| Characteristic | N (%) of participants |
| --- | --- |
| Total survey respondents | 29 |
| Staff physician | 23/29 (79%) |
| Trainee | 6/29 (21%) |
| Parental status |  |
| Childbearing parent | 22/29 (76%) |
| Non-childbearing parent | 7/29 (24%) |
| Number of children |  |
| One child | 19/29 (66%) |
| Two or more children | 10/29 (34%) |
| Stage of career when first child was born |  |
| Staff | 14/29 (48%) |
| Trainee | 15/29 (52%) |
| Post-graduate years 1-3 | 7/15 (47%) |
| Post-graduate years 4+ | 8/15 (53%) |

### Parental leave and coverage

The median duration of parental leave among survey respondents was 5 months (range 1-12 months) (Table 2). Childbearing parents took longer leaves than their non-childbearing counterparts (median 6 months versus 1 month). Factors that influenced decision around length of leave taken are depicted in Figure 1. Finances were a prominent consideration for both staff (64%, 9/14) and trainees (27%, 4/15) at the time when first child was born. Respondents reported not receiving assistance from the organization in arranging coverage for clinical and/or academic work during their leave. Approximately one third of respondents (34%, 10/29) reported feeling pressured to continue work-related activities while on leave. Some described feeling obligated by prior commitments, while others noted feeling pressured by potential loss of career opportunities.

**Table 2.**
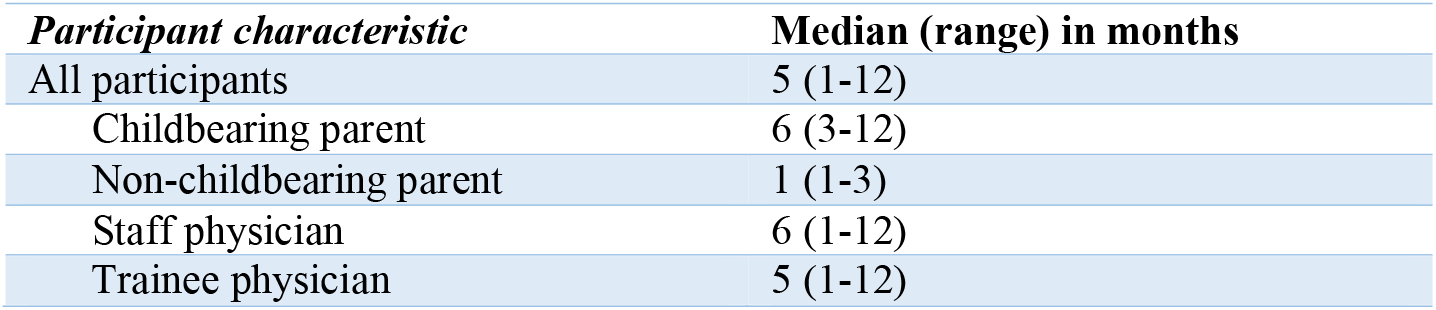
Duration of parental leave by childbearing status and stage of career.

| Participant characteristic | Median (range) in months |
| --- | --- |
| All participants | 5 (1-12) |
| Childbearing parent | 6 (3-12) |
| Non-childbearing parent | 1 (1-3) |
| Staff physician | 6 (1-12) |
| Trainee physician | 5 (1-12) |

**Figure 1.**
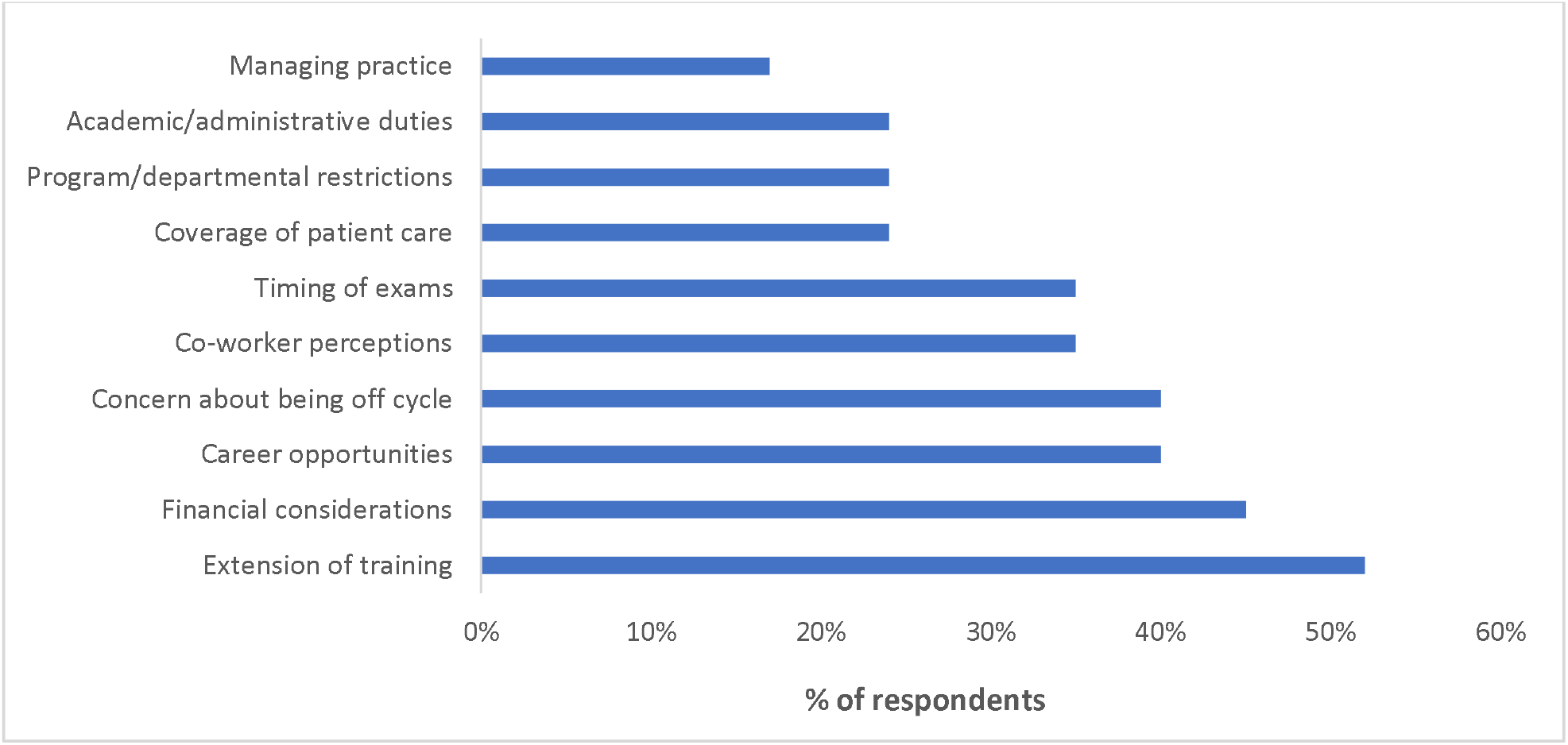
Factors influencing duration of parental leave among study participants.

### Returning to work

Forty-five percent (45%, 13/29) of respondents reported that they were able to return to work in a graded manner. Of those that did not have this option, 56% (9/16) were trainees. Frequently cited facilitators to returning to work included starting with an easier rotation (for residents), fewer call shifts, lighter daytime schedule or part-time work. Approximately half of the staff respondents (at the time of first child born) (43%, *n*=6/14) reported that they had to “pay back” missed time or call shifts, which made the transition back to work more challenging.

> *“I found [paying back time] made my return harder, as I was pumping and caring for small children [at home]*.*”*

### Lactation supports

Among those for whom lactation was applicable (21 of 29 respondents), 76% (16/21) reported that they continued their lactation efforts on returning to work and 24% (5/21) chose not to. Methods of support for lactation included: access to a private room to pump, dedicated time to pump, and access to adequate breastmilk storage space. The quality of these supports, when available, was variable (Figure 2). Five (5/21) participants (24%) noted that they either did not pump at all or quit prematurely due to workplace challenges. Some participants described these issues further:

> *“I regularly pumped in a bathroom stall on the toilet*.*”*
>
> *“[I had] constant worry about patient care during pumping and pager going off*.*”*

**Figure 2.**
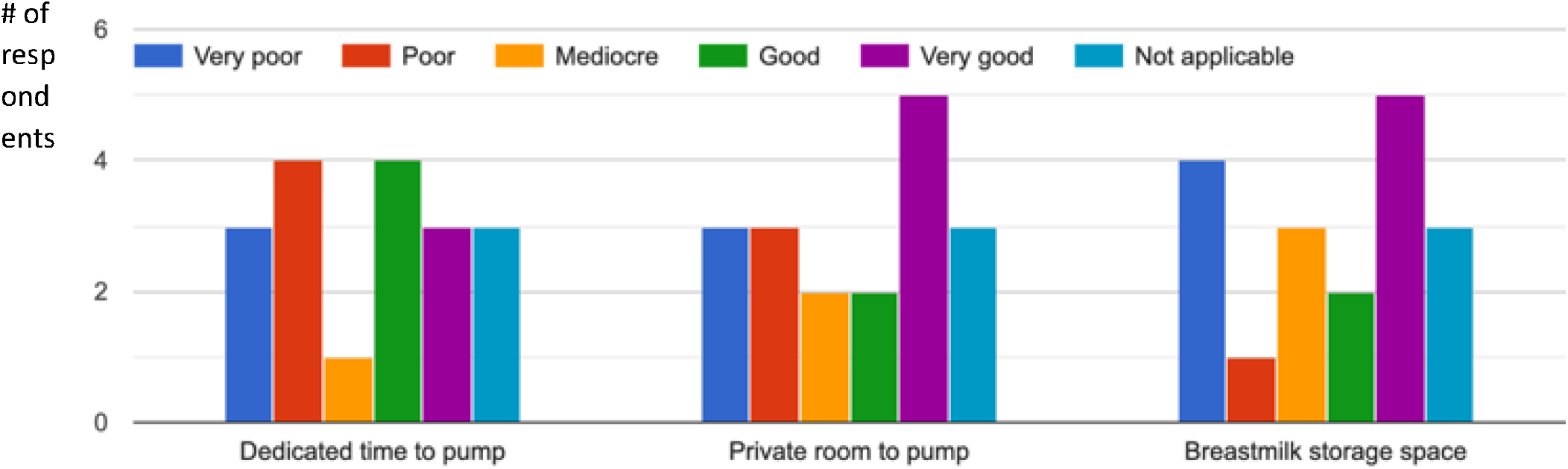
Quality of lactation supports accessible to participants at the workplace.

Approximately 29% of respondents (5/17) reported that they were very uncomfortable or uncomfortable discussing their lactation support needs at the workplace.

### Accommodations for childcare

More than half (59%, 17/29) the respondents reported instances of disruption in their childcare arrangements. Sixty-one percent (63%, *n*=14/22) claimed that they felt supported by leadership in dealing with the ensuing challenges. Of those, 32% (*n*=7/22) felt somewhat supported and 5% (*n*=1/22) felt unsupported. Thirty five percent (35%, *n*=6/17) of respondents cited that they were not provided with duty hour modifications when required; for example, to look after a sick child at home. Some participants described a cultural pressure to remain at work, while one participant described experiences of maternal discrimination:

> *“I find it difficult to have to call in for back up call to stay home to look after child. On return to work (often exhausted from looking after sick child) I am faced to having to make up for the call…*.*”*
>
> “*I didn’t want to get special exceptions and I wanted to finish my residency without drawing attention to myself*.*”*
>
> *“… Males do not get it. … They don’t appreciate all the things we do and that we can’t make it to 7:30 am meetings or might be a little tired because of the 4am fevers. I have specifically heard males in the DOM say, ‘Childbearing women are a liability’*.*”*

### Co-workers

Ninety percent (90%, *n*=26/29) of respondents felt comfortable discussing their dual role of physician-parent with their co-workers. The remaining respondents described feeling that personal life discussions were not appropriate in the workplace.

> *“… work trumps everything. Never felt like anyone cared what anyone had going on outside of work*.*”*
>
> *“I rarely told people I had a child at home so they wouldn’t think I was less of a resident*.*”*

Many respondents (46%, *n*=16/28) felt guilty about burdening their co-workers for requesting accommodations to deal with emergency childcare issues.

> *“I’ve had to leave early a few times and always feel like I’m not ‘pulling my weight’ when I do*.*”*

### Work-life balance

When asked to rate their experiences of balancing their personal and professional lives on a scale of 1-10 (1 being “impossible to maintain” and 10 being “extremely easy to maintain”), most respondents (66%, *n*=19/29) chose a rating of 5 or less (mean rating of 4 among trainees and 5 among staff). Reliable childcare and lighter workload were described as facilitators of work-life balance, while working outside of business hours was reported as a barrier. Other participants described feelings of overwhelm when juggling their work and life responsibilities.

> *“I find that I’m forced to choose between being the resident that I would want to be in an ideal world and time with my child “*

## DISCUSSION

This study supports previous findings that physician-parents face a plethora of challenges integrating their roles in medicine and parenthood, including but not limited to suboptimal duration of parental leave, inadequate lactation supports at the workplace, and inability to secure duty hour modifications for childcare emergencies.

Participants in this study were found to take shorter parental leaves (median 5 months) compared to mothers in the general population, who take about 10 months, according to a Canadian census-based study^15^. Suboptimal childbearing leaves in medicine have been reported previously^16,17^ despite legislation allowing for longer leaves^18^ and evidence of the benefits of parent-child bonding during early infancy^19,20^. In our study, financial constraints, particularly among staff physicians, were a determinant of leave length. This may, in part, be related to a lack of income support after the first 17 weeks of leave, during which time Ontario physicians practicing as independent contractors receive a maximum remuneration of $1,300 weekly through the Pregnancy and Parental Leave Benefit Program in Ontario^21^. As such, medical institutions may consider methods of bolstering parental leave benefits for staff to empower longer leaves, although there is a gap in the current literature regarding how to operationalize or fund such programs. It is noteworthy, however, that paid leave in clinical medicine may lead to downstream cost savings due to increased retention of female staff^22^. Some staff respondents in our study did not receive institutional support in arranging clinical coverage, which was another determinant of leave duration. A previous study identified that only 20% of parental leaves in Ontario were replaced by temporary faculty, although majority of respondents favored replacement and felt it should be institutionally funded^10^. These findings emphasize the need for systems-level oversight in arranging clinical and/or academic coverage, provision of financial incentives for colleagues assuming additional responsibilities, and hiring of additional staff^23-25^. We found that resident physicians were curtailing their parental leaves for distinctive reasons, such as anxiety about extending training and delaying licensure, analogous to findings in previous studies^8,25^. As such, post-graduate medical education programs may consider incorporating flexible graduation requirements, while specialty boards may consider expanding the board testing dates for off-cycle trainees^23,26^.

A significant proportion of respondents in our study reported feeling pressured to continue working during parental leave. Previous research similarly indicates that women in academia engage in career-related activities while on maternity leave to avoid negative career effects from decreased productivity^27,28^. These findings underscore the need for medical institutions to ensure that leaves for family building are not detrimental to future career opportunities through measures like protection of pre-leave academic, administrative, and clinical roles; provision of institutional supports for academic coverage; creation of flexible promotional criteria; and targeted leadership development programs^23,24,29,30^. Many respondents in our study reported that they were not offered an opportunity to transition back to work in a graded manner following parental leave. A phased return is important, especially for resident physicians who are already disadvantaged by an inherent lack of autonomy and fear of authority within the overtly hierarchical structure of medicine^31,32^. Medical education leadership could support residents’ autonomy in their return to work post-leave by considering childcare obligations when scheduling rotations and call shifts, creating parenting-friendly electives, and/or permit the application of credits from administrative or research roles towards program completion^25,33,34^. Moreover, flexible, part-time, and prolonged options for completing training may be considered, as these have been identified as factors enabling retention rates in training programs, career satisfaction, and professional/personal life balance among physician-mothers^23,35,36^.

On return to work, most respondents in our study received accommodations for childcare when requested; however, there was significant stress among participants around burdening colleagues. Clearly delineated family leave policies could destigmatize and streamline the process of requesting accommodation while ensuring institutionally protected time for childcare^23^. Moreover, access to quality, reliable, on-site childcare services and/or financial support to offset the cost of childcare may mitigate childcare emergencies and have been identified as enablers of career satisfaction and successful completion of training programs^23,37^. Our study also found that there were significant shortcomings in the availability and quality of lactation supports offered to physicians, as a result of which some participants endorsed prematurely weaning their lactation efforts. These findings are in keeping with previous research showing that physicians have low breastfeeding maintenance rates compared to the general population^38^. It is, therefore, incumbent on our medical institutions to support lactation by enacting policies to protect time to express milk; providing private, clean, and comfortable lactation rooms, as well as equipment and supplies (such as hospital-grade electric pumps); and promoting lactation awareness at the workplace^5,23^.

We found that co-worker perceptions were factored into decisions about length of leave, and some participants did not feel comfortable asking for accommodation or discussing their parenting role at work due fear of discrimination. These findings are in keeping with previous discussions of negative attitudes towards physician-parents among colleagues and peers^7,30,37^ and highlight the imperative to address gender-based discrimination and enhance workplace acceptability of parenthood in medicine. Specific interventions such as ensuring backup support for unscheduled absences and pathways to report and address harassment, as well as broader strategies to develop strong leadership, role modelling and advocacy in medicine, have been suggested as solutions to workplace discrimination of parents^7,23^. Work-life balance among participants in this study was generally rated as suboptimal. High levels of burnout among physicians, and especially female physicians who often have a disproportionate burden of domestic responsibilities^39^, has been extensively described^40-42^. Burnout has been associated with increasing rates of attrition from the medical profession, as well as depression and death by suicide among physicians. In one survey, residents had the highest level of career dissatisfaction^43^, with feelings of guilt around partner and family neglect due to the demanding residency workload^3^. Targeted interventions to improve work-life integration and mitigate burnout for physician-parents are warranted. Strategies could include experienced female mentorship, facilitated support groups, mental health services, provision of training and career development resources, increased flexibility in work hours and format, and part-time career tracks^7,16,24,25^.

This study has several limitations. First, there was a low response rates despite broad advertisement of the study; however, this is typical of surveys among medical professionals^44,45^. Moreover, the sample population was potentially biased by listservs that allowed circulation and snowball method of recruitment. Second, we only surveyed members of the academic internal medicine community; thus, this study may not be generalizable to the experiences of those who work in non-academic and community settings. Lastly, while the study survey was developed based on a literature review and the experiences of the investigators, its reliability was not formally tested.

## CONCLUSIONS

The findings of this study add to the growing body of research describing the challenges faced by physician-parents at individual, interpersonal, and institutional levels. Our findings re-iterate the need to exact a change in culture to support and normalize parenthood for physicians, which in turn will foster productivity in academic medicine, as wellness and creativity go hand-in-hand^46-48^. Policies and procedures should be revisited using principles of implementation science to ensure that interventions and changes are appropriately mapped to local barriers and facilitators, and are formulated to address systemic inequities and promote career success of physician-parents.

### Ethics approval

The study was approved by the Unity Health Toronto Research Ethics Board in accordance with the Tri-Council Policy Statement Ethical Conduct for Research Involving Humans and ICH Good Clinical Practice Guidelines (REB#22-094).

## Data Availability

All data produced in the present study are available upon reasonable request to the authors

## Authors’ contributions

SH, MF, and MS conceptualized the study. SH collected the data, and drafted the manuscript. All authors critically reviewed the manuscript, contributed to revisions, and read and approved the final manuscript.

## Acknowledgements

We would like to express profound gratitude to Dr. Brittany Lostracco for their careful review of the final version of this manuscript and for the opportunity to include as senior co-author Dr. Michael Fralick, our cherished friend and colleague, who very meaningfully contributed to this work since its inception.

